# Exploring the Relationship Between Non-Suicidal Self-Injury and Problematic Sexual Behaviour

**DOI:** 10.64898/2026.04.17.26351044

**Authors:** Shui Jiang, Jerome C. Foo, Leslie Roper, Esther Yang, Bradley Green, Randolph Arnau, Behavioural Addictions Studies and Insights Consortium, Rohit J. Lodhi, Rick Isenberg, David Wishart, Esther Fujiwara, Patrick J. Carnes, Katherine J. Aitchison

## Abstract

**Objectives:** Non-suicidal self-injury (NSSI) and self-harming sexual behaviours share functional and behavioural overlaps. However, the relationship between NSSI and problematic sexual behaviour (PSB) remains underexplored. This study aimed to investigate the association between NSSI and PSB in two cohorts - a non-clinical university cohort and a clinical PSB patient cohort.

**Methods:** Data were collected from 2,189 university participants and 477 clinical PSB patients. NSSI was assessed via self-report, and PSB was measured with the Sexual Addiction Screening Test-Revised (SAST-R) Core. The four core addictive dimensions of PSB: relationship disturbance, loss of control, preoccupation, and affect disturbance, were also evaluated. Logistic regression analyses were conducted to examine the association between PSB (presence/absence and severity) and NSSI, looking at effects of gender and contributions of addictive dimensions of PSB.

**Results:** Rates of NSSI were similar in the university (7.1%) and patient (5.7%) cohorts; stratified by gender, a higher proportion of women PSB patients had NSSI compared to in the university cohort (29.3% vs 9.3%). In the university group, who had milder PSB than patients, PSB was associated with NSSI (OR=2.11, *p*<0.001); a significant gender by PSB interaction was found showing that women with PSB were over four times more likely to have NSSI than men without PSB (OR=4.44, *p*=0.037). In contrast, PSB severity was not associated with NSSI in PSB patients (OR=1.10, *p*=0.25). Associations of the addictive dimensions of PSB with NSSI were observed only in the subgroup of university women, in the ‘preoccupation’ dimension (*p*<0.001).

**Conclusions:** Our findings highlight gender-specific patterns in the association between PSB and NSSI, suggesting the need for further research and possibly targeted prevention and intervention strategies in women.

## INTRODUCTION

Non-suicidal self-injury (NSSI) is defined as the direct and deliberate destruction of the body in the absence of suicidal intent.(1) Over time, various terms have been used to describe NSSI,(2) including parasuicide,(3) deliberate self-harm,(4) self-mutilation(5) and self-injurious behaviour.(6) Although occasionally conflated with suicidal behaviour, NSSI is distinct due to the absence of suicidal intent.(7) The prevalence of NSSI varies across age groups and populations. A meta-analysis of a non-clinical cohort estimated lifetime NSSI prevalence at 5.5% in adults (aged over 25 years), 13.4% in young adults (18-24 years), and 17.2% in adolescents (10-17 years).(8) Community-based studies suggest a lifetime prevalence of 18-22% in adolescents,(9) while rates in clinical populations range widely (13-80%).(10) Established risk factors for NSSI include younger age,(8, 11) history of childhood trauma,(11) and being of female gender.(12) Multiple theoretical models have been proposed to explain NSSI, emphasizing emotion regulation deficits, psychological distress, and maladaptive coping as key contributors.(13) NSSI behaviours may range from minor acts, such as scratching or skin picking, to more severe forms, including cutting, carving, or bone-breaking.(10, 14) Of note, NSSI may involve harm to intimate areas, such as the breasts or genitals,(15) or manifest as “self-injury by proxy,” where harm is incited by a partner.(10) These behaviours share functional and psychological similarities with problematic sexual behaviour (PSB), suggesting potential links between the two.(16–20)

PSB, also referred to as sex addiction or compulsive sexual behaviour, is characterized by recurrent and uncontrollable engagement in sexual activities that result in adverse social and functional consequences.(21) It is estimated to affect 3-6% or more of the general population.^4^ PSB is commonly conceptualized through four core addictive dimensions: relationship disturbance (sexual behaviour that disrupts interpersonal relationships), loss of control (inability to cease the behaviour despite negative outcomes), preoccupation (persistent obsessive thoughts about sexual behaviour or fantasies), and affect disturbance (emotional distress, such as anxiety or depression, associated with sexual behaviour).(22) Prior research suggested that NSSI and PSB share several addictive-like features, such as loss of control, preoccupation, compulsivity, and tolerance, as well as common underlying processes like maladaptive coping strategies and emotional dysregulation. (21, 23–27)

While sexual behaviour used explicitly for self-harm purpose (i.e., sex-as-self-harm) has been documented primarily in Northern European studies, these studies are either qualitative or case-based research, typically focusing on individual experiences without reference to a PSB clinical framework.(19) As a result, little is known about how clinically defined PSB relates to NSSI. Furthermore, although PSB is more frequently reported in men,(28) women represent a substantial minority (8-40%) of affected individuals, and their experiences are disproportionately understudied.(29) Understanding whether PSB is associated with NSSI, and whether this risk is gender-specific may help identify high-risk subgroups and inform tailored interventions. Thus, our primary aim in this study was to examine the association between PSB and NSSI in a large university cohort, and in a clinical cohort of PSB patients, also examining gender differences. As a second aim, we explored which of the addictive dimensions of PSB (relationship disturbance, loss of control, preoccupation, affect disturbance) are specifically associated with NSSI. This exploratory analysis may offer insight into which aspects of PSB are most relevant to NSSI and help refine dimension-specific risk profiles for future research.

## METHOD

### Participants

The non-clinical university cohort (n = 2,189) has been previously described elsewhere (21) and included 713 men (32.6%) and 1,476 women (67.4%), with a mean age of 21.9 years (SD = 5.1). During screening, a subset of participants (n = 275) met the population threshold for PSB (score ≥ 6 on the Sexual Addiction Screening Test - Revised (SAST-R) Core). This subgroup is referred to hereafter as the “non-clinical PSB group”. The clinical PSB patient group comprised patients recruited from eight addiction treatment facilities in the United States, including outpatient clinics and residential treatment centers (n = 477). Among these patients, 450 (94.3%) were men and 27 (5.7%) were women, with a mean age of 43.6 years (SD = 13.0).

Informed consent was obtained from all participants prior to their participation. The study was approved by the Quorum Review Institutional Review Board (Seattle, Washington; Protocol Number: 2016-001) and the University of Alberta Research Ethics Board (Protocol Number: Pro00066552).

### Measures

#### NSSI

Self-reported histories of NSSI were collected from both cohorts, while current NSSI was assessed in the university cohort only. Historical NSSI was measured by one question: “Indicate if you have a previous diagnosis of NSSI, such as cutting and burning.” Current NSSI was assessed with a similar item that specified timing: “Indicate if you have a *current* diagnosis of NSSI, such as cutting and burning”. Responses were combined to create a composite NSSI variable, where participants were categorized as “yes” (1) if they reported either a history of or current NSSI, and as “no” (0) if they reported neither.

#### PSB and Addictive Dimensions

PSB was assessed using the 20-item Sexual Addiction Screening Test-Revised (SAST-R) Core. Participants responded to each item in a binary format (yes = 1, no = 0). A cutoff score of 6, based on prior studies in the (non-clinical) general population, was used to identify individuals meeting the threshold of PSB in the university cohort.(22) Four addictive dimensions of PSB were evaluated: 1) relationship disturbance: defined as relational problems caused by sexual behaviours (SAST-R Core items 6, 8, 16, and SAST-R item 26); 2) loss of control: referring to the inability to stop sexual behaviours despite negative consequences (SAST-R Core items 10, 12, 15, and 17); 3) preoccupation: characterized by obsessive thinking about sexual behaviours (SAST-R Core items 3, 18, 19, and 20); and 4) affect disturbance: indicating emotional distress such as depression, despair, or anxiety related to sexual behaviours (SAST-R Core items 4, 5, 11, 13, and 14). For each dimension, participants were categorized as “yes” (1) if they endorsed at least two items within that dimension.(22)

#### Covariates: Childhood Trauma, Age, and Gender

Childhood trauma (experiences of any physical, emotional, and sexual trauma) was assessed through self-report in both cohorts. In the university cohort, participants reported on childhood trauma, while in the clinical PSB cohort, both past and current trauma experiences were assessed. A binary childhood trauma variable was created for both cohorts, coded as “yes” (1) if any of the three trauma types - past or current - were endorsed, and “no” (0) if none were endorsed. Self-reported age was collected and treated as a continuous variable. Self-reported gender was coded as a binary variable (men = 0, women = 1).

### Analyses

All statistical analyses were conducted using STATA, Release 17 (StataCorp LLC., College Station, TX, USA).

#### Descriptive Statistics

Clinical and demographic differences between participants with and without NSSI were examined using Pearson’s χ² test for categorical variables (or Fisher’s exact test when expected cell counts were < 5) and the Mann-Whitney U test for continuous variables. Additionally, a gender-stratified analysis was conducted within the university cohort to assess whether NSSI rates differed between participants with and without PSB. This analysis was not conducted in the clinical cohort due to the limited number of women (n =27). Gender differences in trauma were also examined in both cohorts, using χ² tests.

#### Regression Analysis

In the university cohort (n = 2,189), two logistic regression models were conducted on the presence or absence of NSSI. In Model 1, PSB was entered as a binary independent variable (yes vs. no). Due to the right-skewed distribution of PSB scores, with most participants scoring below the threshold in the university cohort, this binary classification was used to distinguish individuals with or without PSB. In Model 2, the same binary PSB variable was entered, with the addition of an interaction term (gender × PSB) to assess potential gender differences in the relationship between PSB and NSSI. As all patients met the PSB threshold (n = 477, “clinical PSB group”), the continuous PSB score was used to represent symptom severity as the independent variable. Age, gender, and childhood trauma were included as covariates in all models. Inclusion of the interaction term was not feasible in the patient cohort, where all participants had PSB and the number of women was too small for reliable gender-based interaction analysis.

#### PSB Addictive Dimensions

Relationships between specific addictive dimensions of PSB (relationship disturbance, loss of control, preoccupation, and affect disturbance) and NSSI were examined in both cohorts, comparing scores between those with and without NSSI.

In the university cohort, we used logistic regressions probing associations between PSB dimensions and NSSI. These analyses focused on women (n = 1,476) or women who met the population PSB threshold (n = 136) owing to female-specific outcomes when we examined the total PSB results.

In the PSB patients, descriptive analyses were conducted to compare endorsement levels across the four PSB dimensions between participants with and without NSSI, with comparisons based on the total number of “yes” responses within each addictive dimension. Regression analyses were not conducted in this group due to limited variability in PSB (all patients met the threshold) and multicollinearity of the four dimensions; the two-sample Wilcoxon rank-sum (Mann–Whitney U) was used.

## RESULTS

### Descriptive statistics

In the university cohort, individuals with PSB reported NSSI at a significantly higher rate compared to those without PSB (13.1% vs. 6.3%, *p* < 0.001). This difference was significant only in women: women with PSB had a significantly higher rate of NSSI compared to women without PSB (24.3% vs. 7.7%, *p* < 0.001), while no significant difference was observed in men (2.8% vs. 2.2%, *p* = 0.68). In the PSB patient cohort, a higher proportion of women reported NSSI compared to men (29.6% vs. 4.2%, *p* < 0.05).

In the cross-cohort comparison, overall NSSI rates did not significantly differ between PSB patients (5.7%, 27/477) and the university cohort (7.1%, 156/2,189) (p = 0.25; Table 1). However, gender-stratified analyses revealed significant differences. In women, NSSI prevalence was significantly higher in PSB patients compared to university cohort (29.6% vs. 9.3%, *p* < 0.001). In contrast, in men, NSSI rates remained low in both groups, with no significant difference between PSB patients and university cohort (4.2% vs. 2.7%, p = 0.15).

**Table 1.**
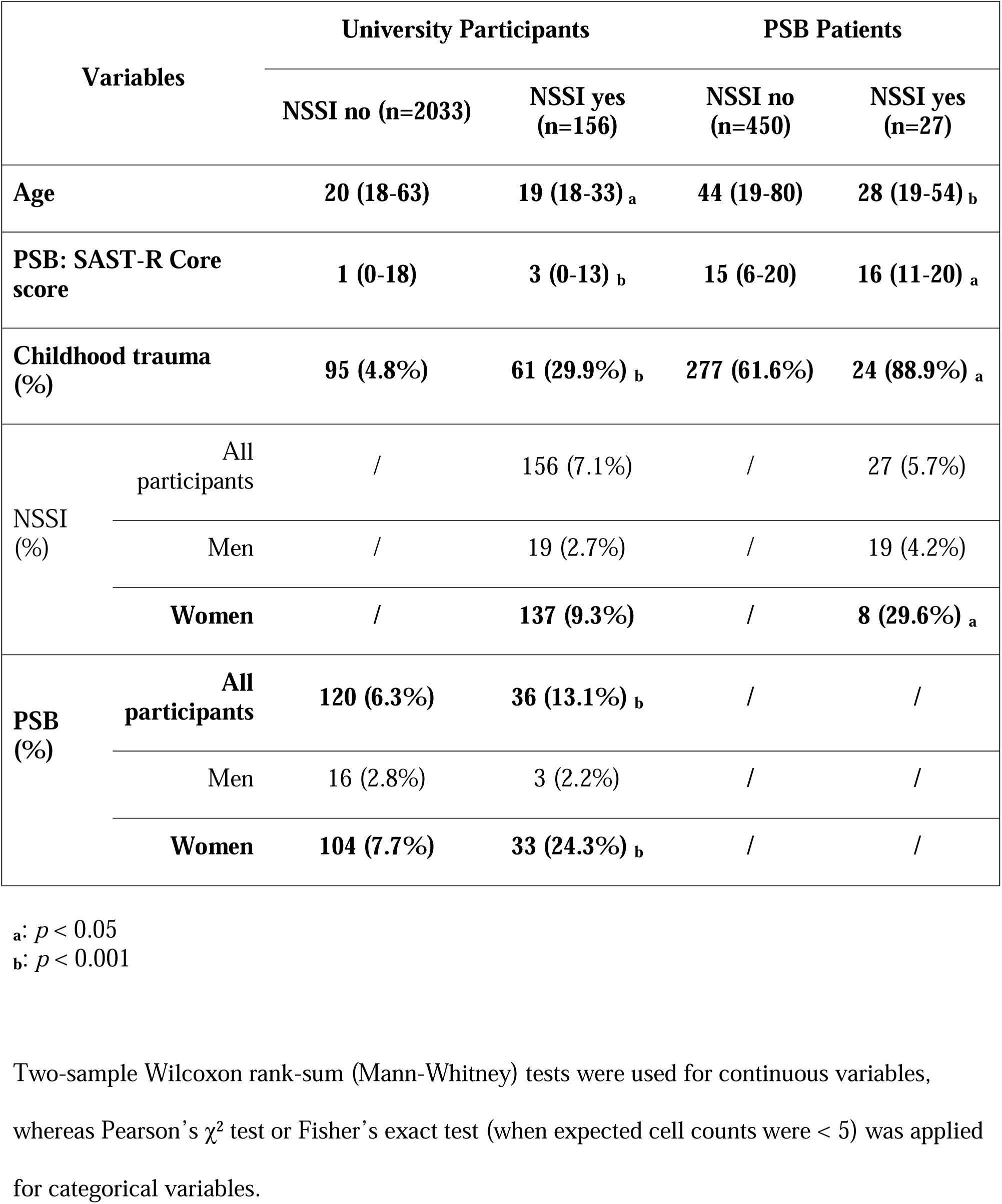
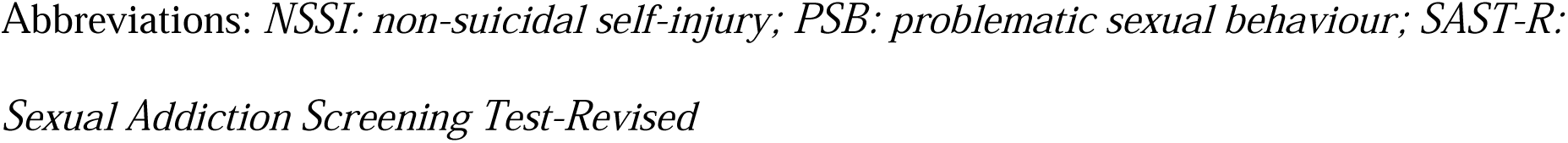
Demographic and Clinical Variables Across Subgroups: Values Are Presented as Medians (Ranges) or Frequencies (Percentages)

In the university cohort, childhood trauma was reported with elevated rates in women (10.6%) compared to men (6.0%, p < 0.001). Childhood trauma was very common in the clinical cohort, again with a significant elevation in women (81.5%) compared to men (62.0%, p = 0.042).

### Regression Analyses

#### University Cohort

Two logistic regression models were conducted to examine the association between PSB and NSSI in university cohort. In Model 1 **(Figure 1)**, PSB was significantly associated with NSSI (OR = 2.11, *p* < 0.001), along with younger age (OR = 0.85, *p* < 0.001), female gender (OR = 3.25, *p* < 0.001), and childhood trauma (OR = 7.61, *p* < 0.001).

**Figure 1.**
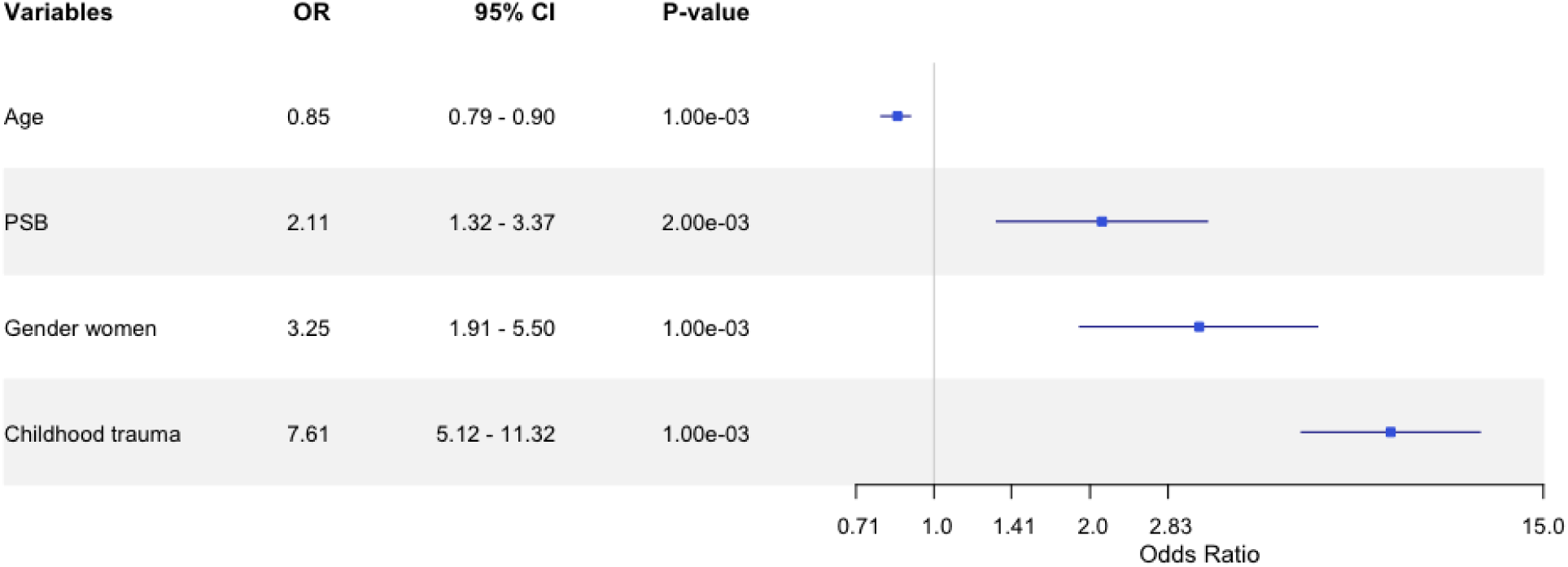
Association Between Problematic Sexual Behaviour (PSB) and Non-Suicidal Self-Injury (NSSI) in the University Cohort. The logistic regression model predicting NSSI demonstrated good fit (n = 2,189), with a log-likelihood of -465.90, pseudo-R² = 0.17, and a statistically significant model overall, χ² (4) = 177.79, *p* < 0.001. Statistically significant predictors of NSSI included younger age (*p* = 1.22 × 10 ), PSB (*p* = 1.93 × 10 ^3^), women gender (*p* = 1.50 × 10 ^5^), and childhood trauma (*p* = 1.12 × 10 ²³).

In Model 2 **(Figure 2)**, younger age (OR: 0.85, *p* < 0.001), female gender (OR: 2.20, p = 0.003), and childhood trauma (OR: 7.76, *p* < 0.001) remained significantly associated with NSSI. Introducing an interaction term between gender and PSB (presence vs. absence) rendered the PSB main effect insignificant (OR = 0.61, *p* = 0.46), but showed that women with PSB had 4.44 times higher odds of NSSI compared to the reference group (men without PSB) (OR = 4.44, *p* = 0.037).

**Figure 2.**
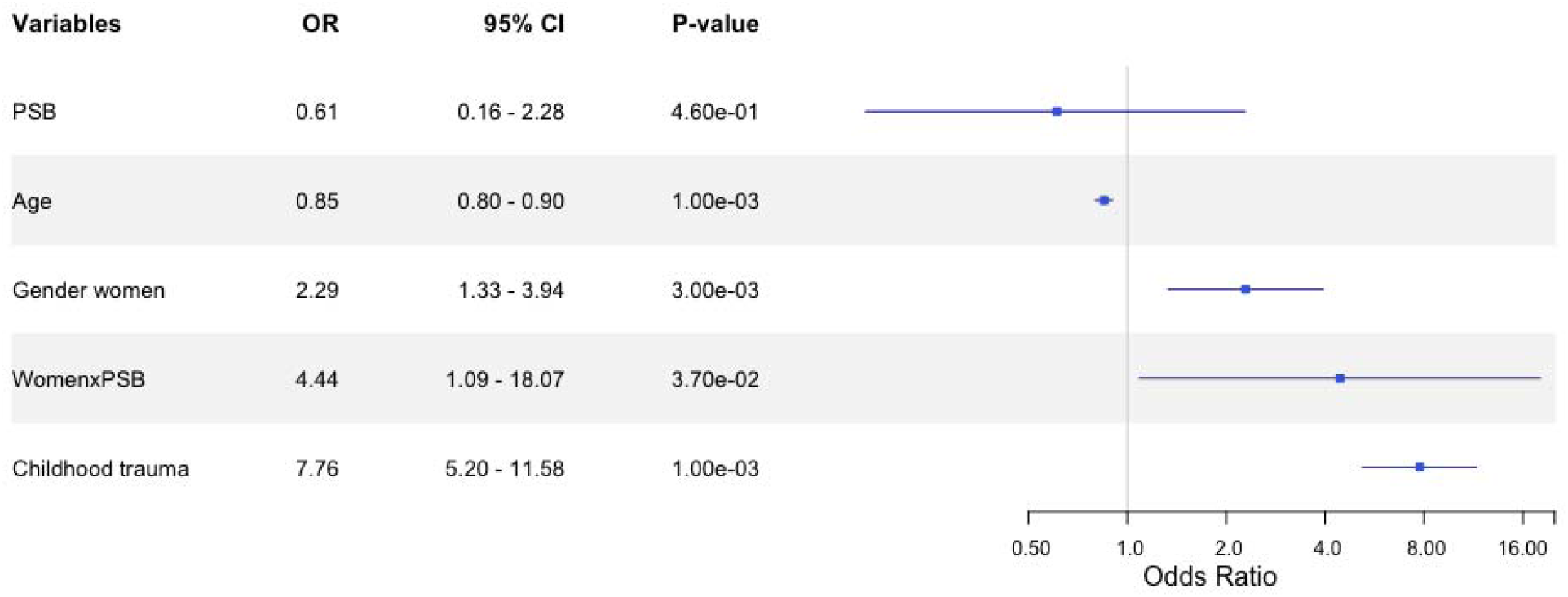
Gender Interaction in the Association Between Problematic Sexual Behaviour (PSB) and Non-Suicidal Self-Injury (NSSI) in the University Cohort. The logistic regression model predicting NSSI demonstrated good fit (n = 2,189), with a log-likelihood of -463.14, pseudo-R² = 0.18, and a statistically significant model overall, χ² (5) = 186.67, *p* < 0.001. Statistically significant predictors of NSSI included younger age (*p* = 2.27 × 10 ), women gender (*p* = 2.67 × 10 ^3^), and childhood trauma (*p* = 1.13 × 10 ²³). The interaction between gender and PSB was also significant, with women who endorsed PSB showing 4.44 times higher odds of NSSI compared to men without PSB (*p* = 0.037).

Women with PSB had the highest predicted probability of NSSI (0.17, *p* < 0.001; 95% CI: 0.11-0.22), followed by women without PSB (0.075, p <0.001, CI: 0.062-0.089), and men without PSB (0.036, p<0.001, CI:0.020-0.053). Men with PSB had the lowest predicted probability of NSSI (0.023, *p* =0.084; 95% CI: -0.0031-0.049). Pairwise comparisons indicated that women with PSB had a significantly higher probability of NSSI than all other groups, including women without PSB (difference = 8.98%, *p* < 0.05), men with PSB (difference = 14.20%, *p* < 0.001), and men without PSB (difference = 12.87%, *p* < 0.001). Furthermore, men and women without PSB differed in NSSI, with higher probability in women (difference = 3.89%, *p* < 0.001), but NSSI did not differ between men with and without PSB (difference = - 1.33%, *p* = 0.40). Thus, women showed higher probability of NSSI than men, with substantially higher rates in women with PSB, but NSSI and PSB appeared unrelated in men.

#### Clinical PSB Patients

In the clinical cohort, younger age (OR: 0.90, *p* < 0.001), female gender (OR:3.64, *p* < 0.001), and childhood trauma (OR:3.79, *p* = 0.013) were significantly associated with NSSI. However, the severity of PSB was not significantly associated with NSSI (OR:1.10, *p* = 0.25, Figure 3).

**Figure 3.**
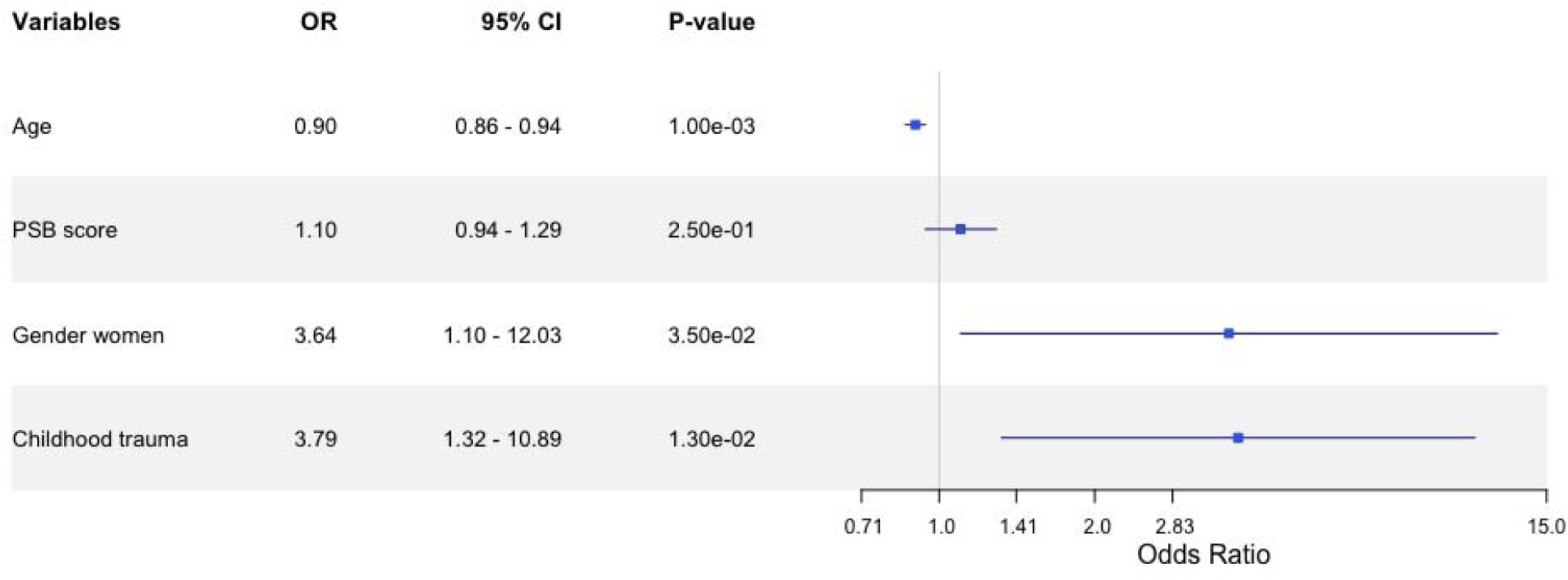
Association Between Problematic Sexual Behaviour (PSB) Scores and Non-Suicidal Self-Injury (NSSI) in the Clinical Cohort. The logistic regression model predicting NSSI demonstrated good fit (n = 477), with a log-likelihood of -78.28, pseudo-R² = 0.25, and a statistically significant model overall, χ² (4) = 46.14, *p* < 0.001. Statistically significant predictors of NSSI included younger age (*p* = 1.96 × 10^D5^), childhood trauma (*p* = 0.035), and women gender (*p* = 0.013).

### Addictive Dimensions

#### University Cohort

We first compared endorsement scores for each PSB dimension between women with and without NSSI. Women with NSSI reported significantly higher scores across all four dimensions: relationship disturbance (*p* < 0.001), loss of control (*p* < 0.001), preoccupation (*p* < 0.001), and affect disturbance (*p* < 0.001). Regression analyses found that the preoccupation dimension was significantly associated with NSSI (OR = 2.69, *p* < 0.001), along with younger age (OR = 0.85, *p* < 0.001) and childhood trauma (OR = 6.90, *p* < 0.001). Relationship disturbance, loss of control, and affect disturbance were not significantly associated with NSSI. This analysis was repeated in women who met the PSB threshold (n = 136). Within this subgroup, women with NSSI reported significantly higher preoccupation scores (*p* < 0.05), but, unexpectedly, lower loss of control scores (*p* < 0.05) compared to those without NSSI.

#### Clinical PSB Patients

In the clinical cohort, individuals with NSSI reported significantly higher endorsement for preoccupation (*p* < 0.05) and affect disturbance (*p* < 0.05) compared to those without NSSI. No significant differences were found for relationship disturbance (*p* = 0.68) or loss of control (*p* = 0.68).

## DISCUSSION

This study provides valuable insights into the relationship between PSB and NSSI, outlining the role of gender and addictive dimensions of PSB. By exploring both a university cohort and clinical patients, we offer a broader understanding of how NSSI and PSB intersect across different groups.

In the university cohort, a significant relationship between PSB and NSSI, particularly in women, was observed. Logistic regression analyses confirmed that women with PSB had a higher likelihood of NSSI. It is important to note that this cohort consisted primarily of young adults (mean age = 21.9), a developmental stage during which NSSI rates typically peak.(2)

Looking at women in clinical PSB patient cohort, the prevalence of NSSI was significantly higher than women in the university cohort (29.3% vs 9.3%). In PSB patients, the severity of PSB was not associated with NSSI, this may be due to cohort characteristics; the clinical cohort was older (mean age = 43.6) and predominantly consisted of men - demographic characteristics that are associated here and elsewhere with lower rates of NSSI.(8, 12) Nevertheless, the small number of female PSB patients showed substantially elevated NSSI rates, suggesting that gender modulates the PSB - NSSI relationship.

Multiple analyses confirmed a strong link between childhood trauma, which was common in both cohorts, and NSSI, highlighting the need for proactive assessment, monitoring and providing support for vulnerable individuals.

### Gender Differences

While our study did not directly examine specific mechanisms, prior research suggests potential reasons that may explain the observed gender differences. In the university cohort, PSB was significantly associated with NSSI in women. Prior research has identified emotional dysregulation and heightened stress sensitivity as characteristics more commonly associated with PSB in women,(30) which may help explain this observed relationship. Furthermore, in our dataset, women in both the university and clinical cohorts reported higher rates of childhood trauma compared to men. A meta-analysis by Calvo et al. (2024) reported strong positive associations between all forms of childhood maltreatment and NSSI across clinical and non-clinical adolescents.(11) This combination of early adversity, emotional dysregulation and stress sensitivity may increase the likelihood of NSSI as a maladaptive coping mechanism in women.(31, 32)

In contrast, apart from internalizing features like depression and anxiety, men with PSB have been reported to have elevated rates of neurodevelopmental psychopathologies such as ADHD and autism spectrum disorder compared to men without PSB, whereas these associations appear weak or absent in women.(30) These factors may contribute to alternative mechanisms, such as impulsivity or externalizing psychopathology, but they may not necessarily lead to NSSI. For example, prior research by Victor et al. (2018) indicated that men are less likely than women to experience urges for NSSI or to use NSSI as a way to regulate emotion.(33) More recent findings further support this difference, suggesting that compared to women, NSSI in men may be more impulsive and associated with externalizing behaviours, such as aggression or substance use.(34) Furthermore, biological factors, including sex-based differences in neuroendocrine responses to stress and emotion regulation,(35) may contribute to divergent patterns of PSB and NSSI across genders. Future research needs to examine how psychological, developmental, and biological factors interact to influence gender- and age-specific patterns in the PSB - NSSI relationship.

### The Role of Addictive Dimensions in PSB and NSSI

Among the PSB addictive dimensions, the most consistent finding across both cohorts was the importance of the preoccupation dimension. In the clinical PSB cohort, individuals with NSSI were more likely to endorse PSB preoccupation than those without NSSI, and in women in the university cohort, preoccupation also emerged as the most consistent PSB dimension related to NSSI. These findings align with research suggesting that obsessive and intrusive sexual thoughts may exacerbate emotional dysregulation,(36) thereby reinforcing the role of preoccupation in maladaptive behaviours such as both PSB and NSSI. Given the cross-sectional nature of the data, these associations should be interpreted cautiously and not assumed to reflect causality. However, these findings highlight the potential value of targeted interventions aimed at reducing preoccupation, such as exposure-based cognitive-behavioural therapy (CBT),(37) which may help lower NSSI risk in those with PSB. Alternatively, as McKay and Andover (2012) suggested, rather than stemming from compulsivity per se, in some individuals, NSSI may be better understood through the lens of rumination and maladaptive emotional avoidance.(38) Since these constructs were not assessed in the current study, our findings cannot directly speak to this possibility, but this perspective highlights the heterogeneous nature of NSSI and the potential utility of personalized treatment approaches that account for differing underlying mechanisms of NSSI.

An interesting finding was that in the university cohort, women meeting the PSB threshold and NSSI reported lower loss of control scores compared to those without NSSI. One possible explanation is that NSSI and PSB may both have emotion regulatory functions. In this context, PSB might be used more deliberately as a way to manage emotional distress, rather than being experienced as uncontrollable or compulsive. This aligns with research in adolescents, which suggests that certain sexual behaviours may be used as a form of self-harm to cope with emotional distress and relational difficulties.(17, 19) Thus, if PSB serves as a coping mechanism to temporality alleviate emotional distress, the need for additional forms of self-harm like NSSI may be reduced.

### Limitations

This study had several limitations. First, the cross-sectional design does not allow to draw causal inferences between PSB and NSSI. Second, the use of self-reported measures could introduce biases, such as response bias, where participants may underreport or overreport sensitive behaviours; and selection bias, as individuals who chose to participate may differ systematically from those who did not, potentially limiting the generalizability of the findings. Additionally, the sample sizes for key subgroups, such as women with NSSI (8/27) in PSB patients, and men with NSSI in both cohorts were smaller, limiting the ability to explore subgroup-specific characteristics and interactions. While this study provides a descriptive understanding of shared features between PSB and NSSI, it does not examine the in-depth psychological, relational, or neurobiological mechanisms underlying these associations.

### Conclusion

This study offers insights into the association between PSB and NSSI in a university cohort and in a clinical PSB cohort. The findings emphasize gender-specific and dimension-specific risks for NSSI. Future research should replicate these findings using matched, randomized samples and, where possible, longitudinal designs to clarify the relationship between PSB and NSSI. Further investigation is needed to confirm the inverse association between NSSI and PSB-related loss of control in high-risk women. Qualitative research may provide additional insights into how addictive dimensions of PSB influence emotion regulation and coping strategies across populations.

## Supporting information

Supplementary Info - BASIC Consortium

## Data Availability

Data produced in the present study may be made available upon reasonable request to the authors.

## Authors’ contribution

RI, RJL, PJC, and KJA contributed to securing funding. RI, RJL, BG, PJC, and KJA contributed to the study concept and design and ethics. RI and PJC oversaw the patient recruitment by the BASIC, supported in terms of data collection by BG and RA, and for sample collection by KJA, and LR. The university participant recruitment was led by KJA, supported by LR and EY. SJ, JCF, DW, EF, and KJA contributed to data analysis, interpretation, statistical analysis, and drafting and revising the paper.

## Ethics

All participants provided written informed consent. The Quorum Review Institutional Review Board (Seattle, Washington; Protocol Number: 2016-001) and the University of Alberta Research Ethics Board (Protocol: Pro00066552) approved the study.

## Acknowledgements

The authors would like to express their gratitude to Dawon Lee for assisting with the study concept, design, and ethics; to Pennie Carnes and Keanna Wallace for sample collection at the university.

