## Supplementary Info - BASIC Consortium for "Exploring the Relationship Between Non-Suicidal Self-Injury and Problematic Sexual Behaviour"

### The Behavioral Addictions Studies and Insights Consortium (BASIC)

Kate Balestrieri, PsyD
Chris Chandler, MA, LMHC
Lauren Dummit, LMFT
Marcus Earle, PhD, LMFT
Greg Futral, PhD
Michelle Gaugh, MA
Piper Grant, PsyD, MPH
Alex Katehakis, PhD, MA, MFT
Barbara Levinson, PhD, RN, LMFT, LSOTP, CSAT Supervisor, CST Diplomate
Andrew Meadows, BS
Dan Morris, LCSW
Isabel Nino-de-Guzman, PhD
Helena Vissing, PsyD

Randolph Arnau, PhD*
Bradley Green, PhD*
Rick Isenberg, MD*
Patrick J. Carnes, PhD*
Katherine J. Aitchison, BM BCh, PhD, FRCPsych*

**lead investigator*

Affiliations

Triune Therapy Group, Los Angeles, CA, USA (KB, LD, HV)
Christian Health Group, La Jolla, CA, USA (CC)
Psychological Counseling Services, Scottsdale, AZ, USA (ME, RI)
Pine Grove Behavioral Health & Addiction Services, Hattiesburg, MS, USA (GF)
Center for Healthy Sex, Los Angeles, CA, USA (PG, AK)
Kavod Center, Rochester, NY, USA (MG, AM, DM)
Center for Healthy Sexuality, Houston, TX, USA (BL)
Gentle Path at the Meadows, Wickenburg, AZ, USA (ING, PJC)
Department of Psychology and Counseling, University of Texas at Tyler, Tyler, TX, USA (BG)
School of Psychology, University of Southern Mississippi, Hattiesburg, MS, USA (RA)
Department of Psychiatry, College of Health Sciences, University of Alberta, Edmonton, AB, Canada (KJA)
Department of Medical Genetics, College of Health Sciences, University of Alberta, Edmonton, AB, Canada (KJA)
Neuroscience and Mental Health Institute, University of Alberta, Edmonton, AB, Canada (KJA)
Women and Children’s Health Research Institute, University of Alberta, Edmonton, AB, Canada (KJA)
Psychiatry Section, Division of Clinical Sciences, Northern Ontario School of Medicine, Thunder Bay, ON, Canada (KJA)
